# Validation of H5 influenza virus subtyping RT-qPCR assay and low prevalence of H5 detection in 2024-2025 influenza virus season

**DOI:** 10.1101/2025.03.17.25324122

**Authors:** David J Bacsik, Margaret G Mills, Luke D Monroe, Cassey Spring, Ailyn C. Perez-Osorio, Ferric C. Fang, Lori Bourassa, Pavitra Roychoudhury, Katharine HD Crawford, Kevin Snekvik, Alexander L Greninger

## Abstract

A sustained outbreak of H5N1 influenza virus among wild fowl and domestic livestock has caused more than 70 zoonotic infections in humans in the United States, including one death. The Centers for Disease Control and Prevention has recommended rapid H5 subtyping for all hospitalized cases with influenza A virus infection to enable prompt initiation of antiviral treatment, as well as infection prevention and implementation of public health measures to control spread. To address these needs, we developed a multiplex RT-qPCR assay to subtype H5 influenza virus in nasal, nasopharyngeal, and conjunctival specimens with a limit of detection of 230 copies/mL. No cross-reactivity was observed with other common respiratory viruses, including seasonal H3N2 and H1N1 influenza A viruses. We retrospectively subtyped 590 influenza A-positive clinical specimens processed by University of Washington labs between March 2024 and February 2025, including 512 specimens collected during the 2024-2025 influenza season, and detected no H5 positives. After clinical implementation, we performed 85 clinically ordered H5 subtyping tests in February 2025 and again detected no positives. This work enhances clinical pandemic preparedness activities and highlights the exceedingly low prevalence of H5N1 influenza virus during the 2024-2025 respiratory season.

**Importance statement:** The spread of H5N1 influenza virus in the United States has led to the culling of almost 200 million birds, infected cow herds across 17 states, and resulted in 70 human infections as of March 2025. Rapid PCR subtyping of H5 influenza virus is critical to inform hospital infection prevention and public health to enable containment of viral transmission. Here, we report the design, validation, and clinical implementation of a multiplex H5-subtyping RT-qPCR assay for nasopharyngeal, nasal, and conjunctival swab specimens. Additionally, we offer the largest reported study of H5 subtyping of influenza A-positive specimens in the United States to date. No H5 infections were detected in 675 samples collected between March 2024 and February 2025 from patients with confirmed influenza A virus infection at a large academic medical center in Seattle, WA.

## Introduction

H5N1 influenza virus, also known as avian influenza, has circulated continuously in North America since 2021 (1). Since late 2023, a sustained outbreak of clade 2.3.4.4b H5N1 influenza A virus has spread among dairy and poultry farms in the United States (2). Currently, two genotypes within clade 2.3.4.4b co-circulate simultaneously: genotype B3.13, which has demonstrated sustained transmission among cattle (2,3), and genotype D1.1, which circulates in wild bird populations and has caused repeated introductions into livestock, including domestic poultry (4) and dairy cattle (5).

Human infections of both genotypes have been detected, with the first outbreak-related case identified in April 2024 (6). To date, more than 70 cases have been reported in North America (7). Although most cases have not required hospitalization, two cases of critical illness have been detected, including one death in Louisiana (8,9). Conjunctivitis has been the most common symptom reported (10). Exposure to infected livestock is a significant risk factor, and farm workers have been disproportionately impacted (10). In a recent serological survey of dairy workers on H5N1-infected farms, 7% had evidence of recent infection with a subtype H5 influenza virus (11), suggesting that the prevalence of infection in this population could be much higher than has been recognized.

Prompt identification of H5N1 influenza virus infection is necessary to provide appropriate medical care and to contain potential further spread of the virus. The Centers for Disease Control and Prevention (CDC) recommends that all hospitalized patients with influenza A virus infections have subtyping performed within 24 hours of admission–especially patients who are critically ill (12). Because H5N1 influenza virus can cause severe illness, and because circulating strains remain sensitive to antiviral medications, treatment with oseltamivir is indicated for symptomatic patients with suspected H5N1 influenza virus infections (13). Furthermore, airborne precautions and patient isolation are recommended for infection control (14).

In addition to informing clinical management, molecular diagnosis of H5N1 improves influenza surveillance. Though efforts have been made to increase surveillance of non-subtypeable influenza, much of the current surveillance infrastructure has focused on monitoring epidemiologically-linked cases with known exposure to infected animals, and it is possible that cases without recognized risk factors have gone undetected (15). This pattern has occurred during previous outbreaks, including SARS-CoV-2 (16). To provide unbiased monitoring of H5N1 infections among all influenza cases, integration within health systems is required. To increase testing capacity, the CDC has issued a call (17) requesting the development and implementation of laboratory developed tests to detect subtype H5 influenza virus (18–20).

Here, we report our validation of a multiplex RT-qPCR assay for H5 influenza virus subtyping in nasopharyngeal, nasal, and conjunctival swabs. We have used the assay to test 675 influenza A-positive specimens processed by the University of Washington Laboratories between March 2024 and February 2025, detecting no cases of H5 influenza virus to date.

## Materials and Methods

### Clinical specimens and associated metadata

Residual deidentified nasopharyngeal swabs, nasal swabs, and conjunctival swabs from the UW Virology Laboratory were used for validation. Retrospective subtyping was performed by testing available residual influenza A-positive respiratory specimens from UW Medicine with collection dates of March 2024 or later. These dates were chosen based on the date of the first human H5N1 infection reported in the United States (21). Available samples had been previously collected for genomic surveillance and had Ct values less than 31. After assay validation and implementation at UW Medicine, test results and metadata were obtained from the laboratory information system. This study was approved by the UW Institutional Review Board with a consent waiver (STUDY00010205).

### RNA extraction

RNA was extracted from clinical specimens on a Roche MagNA Pure 96 instrument using the Viral NA Small Volume Kit (06543588001). For each sample, 200 µL of specimen were used as input and RNA was eluted in 50 µL of elution buffer. An exogenous RNA template was included with the lysis buffer as an extraction control (Table 1).

**Table 1:**
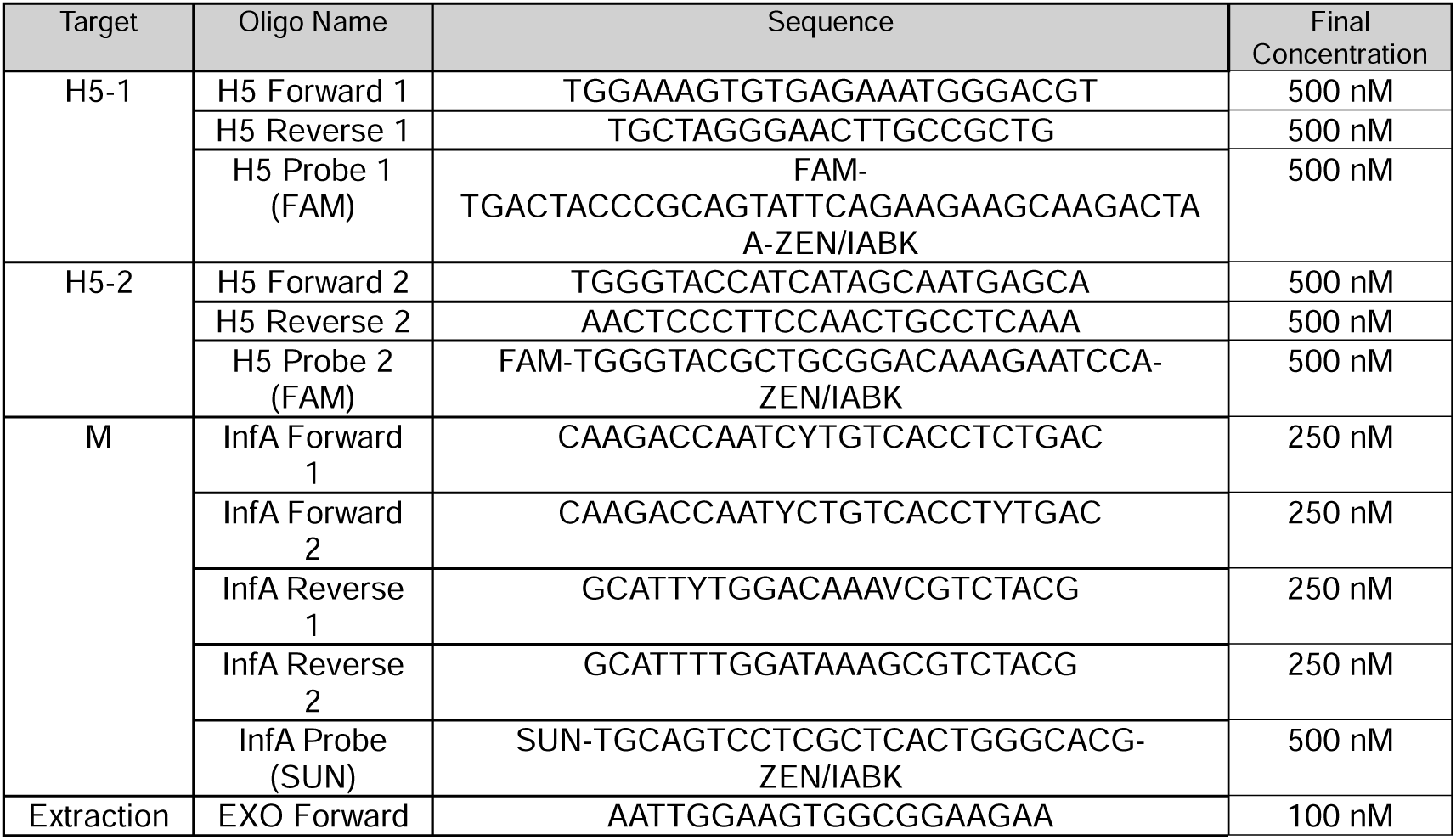
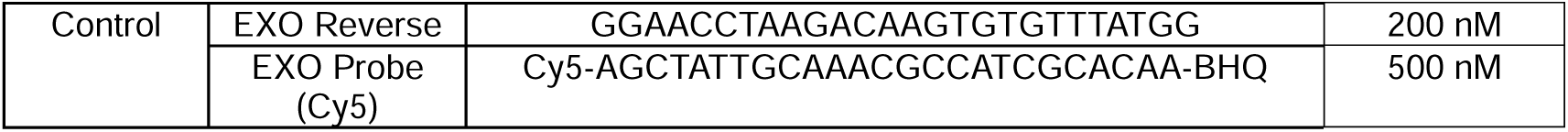
Primer and probe sequences and concentrations.

### H5 templates

Because clinical specimens containing H5N1 influenza virus were not readily available during test validation, we employed four alternative sources of H5 RNA: two sets of synthetic RNA templates, a collection of H5-positive RNA samples extracted from animal specimens, and inactivated H5N1 virus reference material. The first set of synthetic RNAs were transcribed *in vitro*, referred to as “IVT RNA templates.” The H5 RNA was transcribed from a gBlock containing the *HA* coding sequence of the clade 2.3.4.4b H5N1 virus strain A/white-tailed eagle/Hokkaido/20220322001/2022 (Supplementary File 1) using the New England Biosystems HiScribe T7 RNA Synthesis Kit (E2040). The M RNA was prepared as previously described (22). A second set of synthetic RNA was obtained from the National Institute of Standards and Technology (NIST) as H5N1 (Avian Influenza) Synthetic RNA Fragments (Research-Grade Test Material 10263). The NIST RNA set includes synthetic RNAs encoding the *HA*, *M,* and *NA* genes of the clade 2.3.4.4b H5N1 virus strain A/American Wigeon/South Carolina/22/2021 in a background of 5 ng/µL Jurkat cell RNA.

Authentic H5N1-positive RNA samples (n=7) were obtained from the Washington State University Animal Disease Diagnostic Laboratory. The samples were previously extracted from a variety of animal-source specimens, including mammalian respiratory swabs, mammalian tissue, bulk milk, and avian oropharyngeal/ cloacal swabs (Table S1). All were known to be influenza A-positive by prior RT-qPCR and known to contain H5N1 RNA by prior whole genome sequencing. Inactivated H5N1 virus was obtained from BEI Resources (NR-59886), containing tissue-culture adapted strain A/bovine/Ohio/B24OSU-439/2024 and cell lysate that had been inactivated by gamma-irradiation.

### Digital Droplet RT-PCR

The absolute copy number of the IVT RNA and NIST RNA templates was measured by digital droplet RT-PCR (RT-ddPCR). The absolute copy number of the inactivated H5N1 virus, which was sent unitless from BEI, was also quantified following extraction. Digital droplet PCR was performed with the BioRad One-Step RT-ddPCR Advanced Kit for Probes (1864022) using the H5-2 and M primers and probes from the RT-qPCR assay (Table 1). Each 25 µL reaction was prepared by combining 5 µL of extracted RNA with 20 µL of master mix containing 6.25 µL Super Mix, 2.5 µL RT enzyme, 1.25 µL DTT, primers at a final concentration of 900 nM for H5-2 and 450 nM each for M, and probes at a final concentration of 250 nM. 20 µL of the total reaction mix was used for droplet generation using a BioRad Automated Droplet Generator, and 40 µL of droplet suspension was transferred to a 96-well plate. Reactions were run on a BioRad C1000 Touch thermocycler, using the following conditions: 50°C for 60 minutes, 95°C for 10 minutes, 40 cycles of 95°C for 30 seconds followed by 60°C for 1 minute, and 98°C for 10 minutes. Fluorescence was measured on a BioRad QX600 Digital Droplet Reader using the absolute quantification method.

### RT-qPCR reaction conditions

Multiplex RT-qPCR was performed using the AgPath-ID One-Step RT-PCR Kit (AM1005). For each reaction, 5 µL of extracted RNA were added to a reaction mix containing 12.5 µL AgPath Master Mix, 1 µL AgPath Enzyme, and primers and probes at specified concentrations (see Table 1); the total reaction volume was 25 µL. Reactions were run on ABI 7500 Thermocyclers. Cycle parameters were 45°C for 10 min, 95°C for 10 min, and 45 cycles of 95°C for 15 sec followed by 60°C for 45 sec. ROX normalization and automatic baseline were used for all fluorophores. The threshold values were 0.18 for the H5 target (FAM), 0.16 for the influenza A target (VIC), and 0.1 for the extraction control target (Cy5).

### Computational analysis of primer and probe sequences

To assess the similarity between the chosen oligonucleotides and contemporary North American viruses, computational analysis was performed. All H5 HA sequences collected in North America between January 1, 2024 and January 31, 2025 were downloaded from NCBI (23). The sequences were aligned using MAFFT (24). Binding regions were annotated, and each primer or probe was analyzed independently. Sequences that lacked coverage at one or more nucleotides in the binding site were excluded. The number of mismatches was counted; ambiguous nucleotides were conservatively treated as mismatches.

### Limit of detection

The limit of detection of the multiplex RT-qPCR reaction was determined using IVT H5 (stock 1.07×10^6^ copies/µL by RT-ddPCR) and NIST M (1.47×10^5^ copies/µL by RT-ddPCR) RNA templates. Ten microliters of each RNA template were combined and diluted in 80µL of AE buffer. This stock was diluted 10-fold in AE buffer for initial limit of detection determination and 2-fold around the initial limit of detection for confirmation. Twenty replicates of each concentration were tested, and the 95% limit of detection (LOD95) was determined empirically as the lowest concentration with at least 19 of the replicates yielding a positive result.

### Accuracy

Specimen compatibility and assay accuracy were evaluated using samples with known H5 status. Three types of H5-negative specimens were used for assay specificity: nasopharyngeal swabs (n=20), nasal swabs (n=29), and conjunctival swabs (n=18). The respiratory specimens were confirmed as negative for influenza A/B virus by Cepheid Xpert® Xpress CoV-2/Flu/RSV plus or Hologic Panther Fusion Flu A/B/RSV assays. Conjunctival swabs were collected for HSV and/or VZV testing. Specificity was further evaluated using residual RNA samples known to contain common respiratory viruses by clinical testing via Cepheid or Hologic respiratory panel tests or whole genome sequencing. These included influenza A virus subtypes H3N2 (n=21) and H1N1 (n=17), influenza B virus (n=3), respiratory syncytial virus (n=7), and SARS-CoV-2 (n=10).

Assay sensitivity was assessed using two types of H5-positive samples: H5N1-positive RNA samples extracted from animal specimens (n=7) and contrived H5N1-positive clinical specimens spiked with inactivated H5N1 virus (n=69). To prepare contrived specimens, inactivated H5N1 virus stock, which was received unitless from BEI, was added at a ratio of 1:50,000 by volume to the nasal and conjunctival swab VTM, and 1:10,000 to the nasopharyngeal swab VTM. Specimens with at least 220 µL volume were tested individually, while specimens with less volume were pooled and aliquoted. In total, 29 nasal specimens were prepared (20 individual and 9 pooled); 20 conjunctival specimens were prepared (7 individual and 13 pooled); and 20 nasopharyngeal specimens were prepared (20 individual and 0 pooled). Contrived specimens were extracted as described above.

### Comparison to CDC Influenza A/H5 Subtyping Kit

Since only one RT-qPCR assay is FDA-cleared for H5N1 testing, we compared our multiplex RT-qPCR assay with the CDC’s Influenza A/H5 Subtyping Kit (Version 4, #FluIVD03-11). The CDC H5 Subtyping Kit and the novel multiplex assay were run in parallel on the same set of 10-fold serial dilution templates. For the IVT RNA and NIST RNA templates, dilution series were prepared by combining H5 and M templates at a ratio of 1:1 by volume and serially diluting the template mix in AE buffer. The inactivated H5N1 virus was initially diluted 1:100 in PBS, and a 10-fold dilution series in PBS was prepared from this stock with RNA extracted as described above. The absolute copy number of each template stock was determined by RT-ddPCR after dilution (and extraction, in the case of the inactivated virus). The CDC assay was run according to the manufacturer’s protocol using the Invitrogen SuperScript III Platinum One-Step Quantitative RT-PCR System on the ABI 7500 FAST thermocycler. For each singleplex reaction in the CDC kit, 5 µL of nucleic acid template were added to a 20 µL reaction mix containing 12.5 µL of 2X PCR master mix, 5.5 µL water, 0.5 µL enzyme, and 1.5 µL combined primer/probe mix prepared according to the kit protocol; total volume was 25 µL.

## Results

### Design and computational analysis of primer and probe sequences

A three-channel multiplex RT-qPCR reaction was designed. The primary target was the H5 influenza virus *HA* gene. To reduce the risk of false-negative results due to viral evolution (25) or reagent failure (26), primers and probes were selected to target two non-overlapping regions of the gene. One H5 target (H5-1) generated a 149 bp product from positions 1481-1629 of the HA CDS (GenBank accession LC730539; Figure S1). The other H5 target (H5-2) used oligonucleotides designed by Sahoo et al. (18) and adapted from sequences originally designed by the Hong Kong Centre for Health Protection (27). This target generated a 144 bp product from positions 1101-1244 of the HA CDS. As a pan-influenza A virus control, a highly-conserved region of the influenza A *M* gene was targeted using primers and probes designed by the CDC National Center for Immunization and Respiratory Diseases (28). Oligonucleotides targeting an exogenous RNA template encoding the *TMP1* gene from the marine species *Podocornye carnea* were included as an extraction control (29).

The similarity between the oligonucleotide sequences and contemporary North American H5 influenza virus sequences was assessed computationally. A total of 2,788 H5 *HA* sequences collected in North America between January 1, 2024, and January 31, 2025 were retrieved from NCBI GenBank (Table 2). Of the six oligonucleotides targeting H5 *HA*, five matched recent sequences closely, with two or fewer mismatches in 99% of sequences. One oligonucleotide (H5 Probe 1) displayed two mismatches in 92% of recent H5 sequences, and three or more mismatches in 8% of sequences. This probe was designed to bind to a reference material containing inactivated H5N1 virus from 2009 (BEI Resources NR-59421) with a divergent *HA* sequence because materials for validation were highly limited when assay development began. Despite these sequence differences, the probe remains functional with contemporary templates. The same mismatches are present in the IVT RNA target, which is reliably detected by H5 Probe 1 alone (Table S2).

**Table 2:** Similarity of oligonucleotide sequences and recent H5 HA sequences collected in North America. Primer and probe sequences used in the multiplex RT-qPCR assay were computationally compared to recently circulating H5 *HA* sequences available on NCBI.

| Name | Sequences | 0 Mismatch | 1 Mismatch | 2 Mismatches | 3+ Mismatches | 0 Mismatch % | <=2 Mismatch % | 3+ Mismatch % |
| --- | --- | --- | --- | --- | --- | --- | --- | --- |
| **All Sequences** | **2788** |  |  |  |  |  |  |  |
| H5 Forward 1 Primer | 2759 | 2732 | 27 | 0 | 0 | 99.0% | 100.0% | 0.0% |
| H5 Forward 2 Primer | 2782 | 2736 | 38 | 1 | 7 | 98.3% | 99.7% | 0.3% |
| H5 Probe 1 | 2759 | 0 | 3 | 2537 | 219 | 0.0% | 92.1% | 7.9% |
| H5 Probe 2 | 2781 | 2615 | 154 | 0 | 12 | 94.0% | 99.6% | 0.4% |
| H5 Reverse 1 Primer | 2755 | 2589 | 154 | 7 | 5 | 94.0% | 99.8% | 0.2% |
| H5 Reverse 2 Primer | 2780 | 2709 | 55 | 4 | 12 | 97.4% | 99.6% | 0.4% |

### Accuracy

Three types of clinical specimens were evaluated for compatibility with the multiplex RT-qPCR assay (Table 3). Residual nasopharyngeal, nasal, and conjunctival swab specimens known to be influenza A-negative were extracted and tested. All specimens tested negative for both the H5 and M targets, and tested positive for the extraction control. Therefore, no false-positives or PCR inhibition were observed, and all three specimen types are compatible with the assay.

**Table 3:** Specificity and sensitivity of multiplex RT-qPCR assay. Specificity was assessed using influenza-negative specimens and residual RNA containing common respiratory pathogens. Sensitivity was assessed using animal samples containing H5N1 virus genomes and contrived clinical specimens spiked with inactivated virus.

| Sample Type | N | H5 Positive | IAV M | Extraction Control Positive |
| --- | --- | --- | --- | --- |

**Table 3: Specificity and sensitivity of multiplex RT-qPCR assay.** Specificity was assessed using
|  |  |  | Positive |  |
| --- | --- | --- | --- | --- |
| **Influenza-Negative Clinical Specimens** |  |  |  |  |
| Nasopharyngeal Swab | 20 | 0 | 0 | 20 |
| Nasal Swab | 29 | 0 | 0 | 29 |
| Conjunctival Swab | 11 | 0 | 0 | 11 |
| **H5-Negative Clinical Samples** |  |  |  |  |
| Influenza A (H3N2) Virus | 21 | 0 | 21 | 21 |
| Influenza A (H1N1) Virus | 17 | 0 | 17 | 17 |
| Influenza B Virus | 3 | 0 | 0 | 11 |
| Respiratory Syncytial Virus | 7 | 0 | 0 | 7 |
| SARS-CoV-2 Virus | 10 | 0 | 0 | 10 |
| **H5-Positive Samples** |  |  |  |  |
| Animal Samples | 7 | 7 | 6 | 7 |
| Contrived Nasopharyngeal Specimens | 20 | 20 | 20 | 20 |
| Contrived Nasal Specimens | 29 | 29 | 25 | 29 |
| Contrived Conjunctival Specimens | 20 | 19 | 18 | 20 |

To assess the assay’s specificity, residual RNA samples that were known to contain other common respiratory pathogens were tested (Table 3). Importantly, we included non-H5 influenza A virus subtypes H3N2 and H1N1. We also included samples containing influenza B virus, respiratory syncytial virus, and SARS-CoV-2. None of the samples produced a positive result for the H5 target, demonstrating 100% negative agreement with prior clinical testing. The influenza A samples appropriately amplified the M target, while samples containing other pathogens did not.

Finally, to assess the assay’s sensitivity, two sources of H5-positive samples were used. We obtained 7 RNA samples that were known to contain authentic H5N1 RNA by whole genome sequencing (Table S1) and included both genotype B3.13 and genotype D1.1 genomes. To validate the performance of the assay including the extraction step, we also generated contrived H5-positive samples by adding inactivated H5N1 virus to influenza-negative nasopharyngeal, nasal, and conjunctival swab specimens.

All 7 animal samples amplified the H5 target, demonstrating 100% positive agreement. One sample failed to amplify the pan-influenza A virus M target; this sample had a late H5 Ct value of 38, suggesting a low total amount of viral material. This low-positive specimen indicates that the H5 target was more sensitive than the M target. Following extraction and RT-qPCR testing of the contrived positive specimens, all 20 nasopharyngeal swab specimens and all 29 nasal swab specimens tested positive for the H5 target, demonstrating 100% positive agreement. Of the conjunctival swabs, 19 of 20 samples tested positive for the H5 target, demonstrating 95% positive agreement. Combining all true positive and true negative results, the accuracy for the H5 target was 99.5% and the accuracy for the M target was 96.4% (Table 4).

**Table 4:** Accuracy of influenza virus targets. Samples with known influenza status and known H5 status were used for validation. Due to the lack of H5-positive clinical samples available for validation, a combination of animal-source RNA samples and contrived specimens was used for H5-positive samples.

|  |  | H5-Negative<br>Clinical Samples |  | H5-Positive<br>Animal Samples |  | Contrived H5-Positive<br>Clinical Specimens |  |
| --- | --- | --- | --- | --- | --- | --- | --- |
|  |  | Test<br>Positive | Test<br>Negative | Test<br>Positive | Test<br>Negative | Test Positive | Test Negative |
| H5 Target | Known<br>Positive | 0 | 0 | 7 | 0 | 68 | 1 |
|  | Known<br>Negative | 0 | 118 | 0 | 0 | 0 | 0 |
| M Target | Known<br>Positive | 38 | 0 | 6 | 1 | 63 | 6 |
|  | Known<br>Negative | 0 | 80 | 0 | 0 | 0 | 0 |

### Limit of detection

The analytical sensitivity of the assay was measured using a 2-fold serial dilution of H5 and M IVT RNA templates (Table 5). The 95% limit of detection (LOD95) was determined empirically. The LOD95 for the H5 target was 4.7 copies per reaction, corresponding to 230 copies/mL. The LOD95 for the M target was 36.3 copies per reaction, corresponding to 1800 copies/mL.

**Table 5:** Limit of detection. N = 20. Targeted copies per reaction is based on absolute concentration of stock measured by RT-ddPCR.

| Target |  |  |  |  |  |
| --- | --- | --- | --- | --- | --- |
| H5 | Targeted<br>Copies/rxn | 4.7 | 2.4 | 1.2 | 0.6 |
|  | Avg. Ct | 34.6 | 37.4 | 38.2 | 39.0 |
|  | Std. Dev. | 1.1 | 2.2 | 1.9 | 0.9 |
|  | % CV | 3.3% | 5.8% | 5.0% | 2.3% |
|  | # Detected | 20 | 18 | 15 | 8 |
| M | Targeted Copies/rxn | 72.5 | 36.3 | 18.1 | 9.1 |
|  | Avg. Ct | 35.4 | 36.3 | 38.7 | 40.2 |
|  | Std. Dev. | 0.8 | 1.1 | 1.2 | 1.8 |
|  | % CV | 2.2% | 3.1% | 3.0% | 4.4% |
|  | # Detected | 20 | 19 | 17 | 10 |

### Comparison to CDC Influenza A/H5 Subtyping Kit

The assay’s performance was benchmarked against the CDC’s Influenza A/H5 Subtyping Kit to determine its suitability for surveillance and clinical applications (Figure 1). Ten-fold serial dilutions of the IVT RNA, NIST RNA, and inactivated virus templates were prepared, and the same aliquots were used for testing in parallel. In general, the CDC H5 Subtyping kit produced lower Ct values than the multiplex assay. At the lower extreme of template concentration, the CDC H5 Subtyping Kit detected several samples that were not detected by the multiplex assay, suggesting higher sensitivity. Above concentrations of 10 copies per reaction, corresponding to 500 copies per milliliter, performance was similar between the single-target reactions of the CDC kit and our multiplex assay.

**Figure 1:**
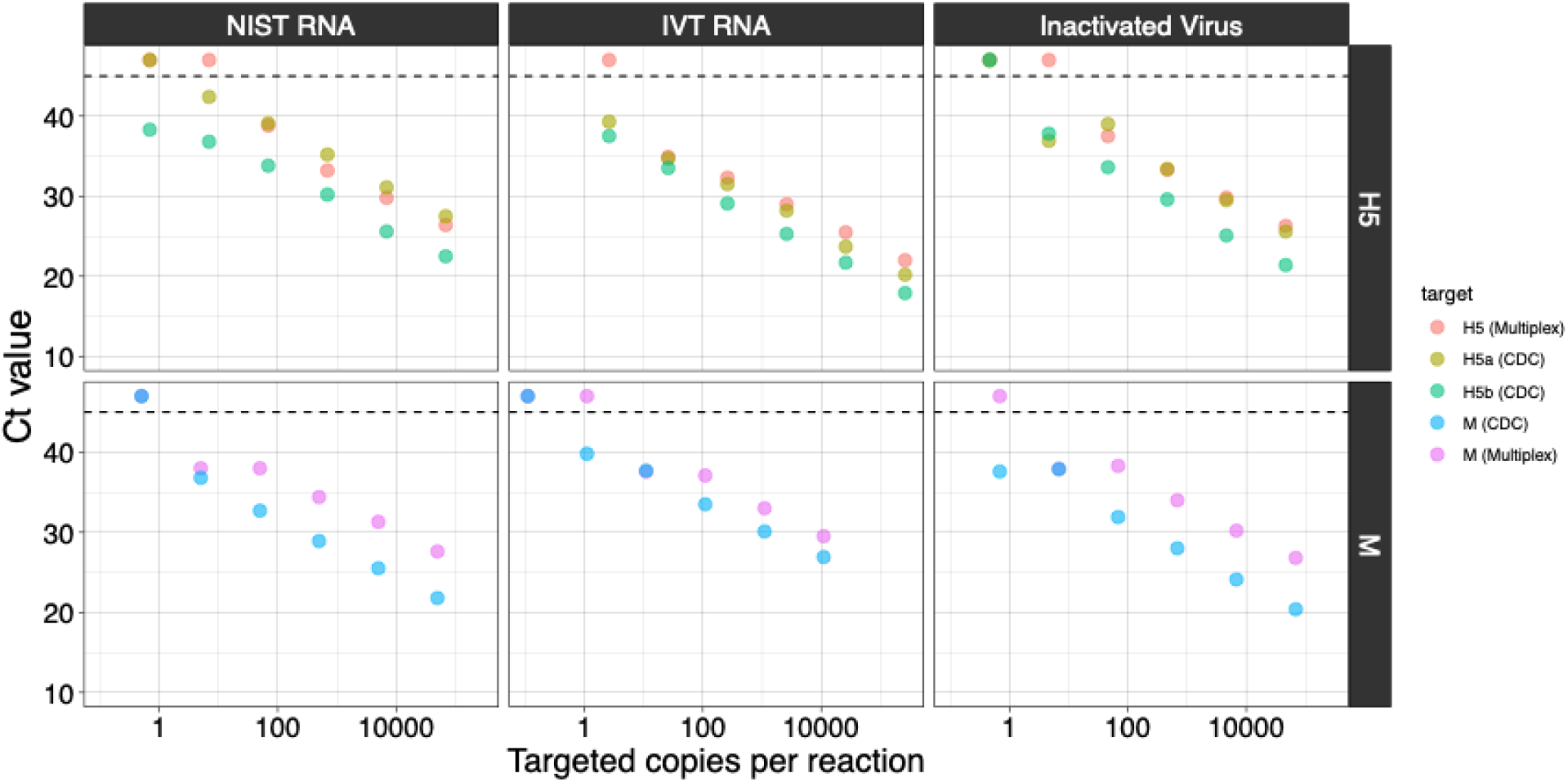
Comparison of multiplex H5 assay and CDC Influenza A/H5 Subtyping Assay. Targeted absolute copy numbers based on RT-ddPCR measurements are shown on the X-axis. Points appearing above the dashed line were not detected after 45 cycles of PCR. Average Ct of two replicates is displayed.

**Figure 2:**
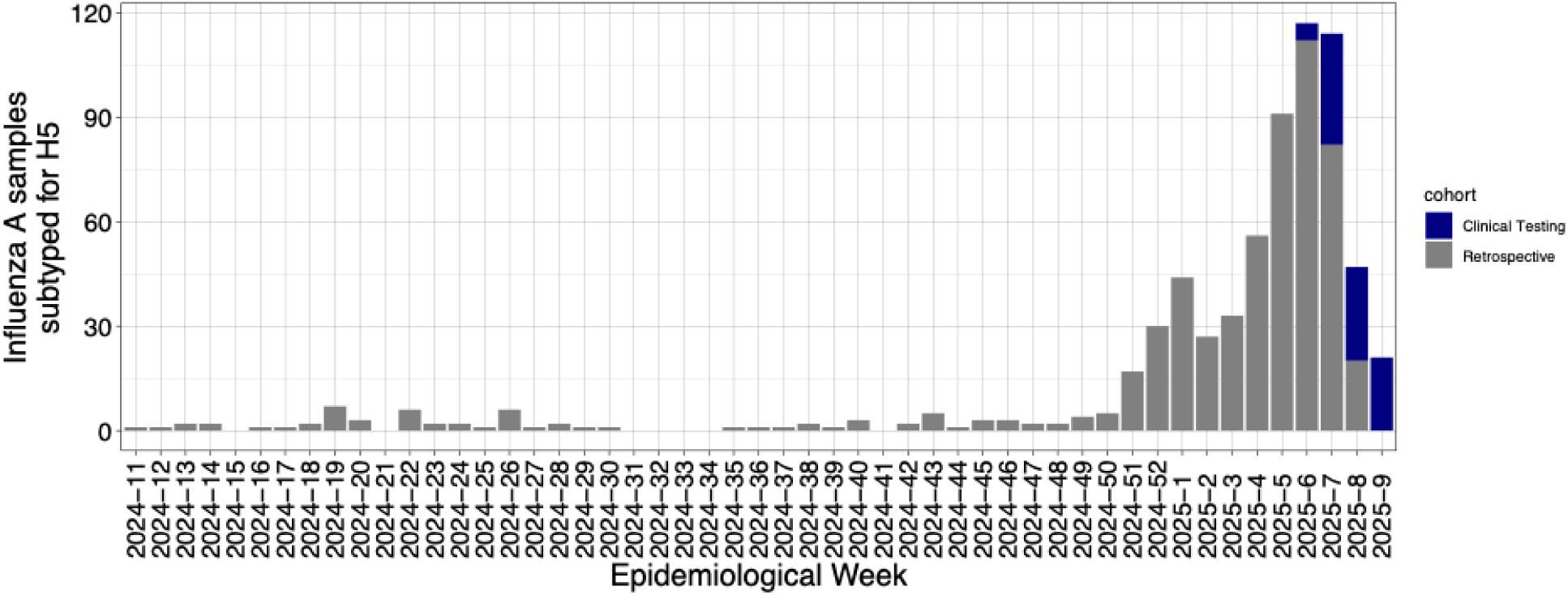
Influenza A-positive samples subtyped for H5 by epidemiological week. Grey bars indicate samples collected for influenza virus surveillance that were retrospectively subtyped for H5 by multiplex assay. Blue bars indicate samples that were ordered for clinical testing after implementation at UW Medicine.

### Retrospective subtyping of influenza A-positive samples collected March 2024 - February 2025

We retrospectively tested 590 residual samples that had been collected for influenza virus genomic surveillance between March 11, 2024 and February 18, 2025. These specimens tested positive for influenza A virus with a Ct value less than 31 on Cepheid Xpert® Xpress CoV-2/Flu/RSV plus or Hologic Panther Fusion Flu A/B/RSV assays at UW Medicine labs. The mean Ct value of samples with a positive influenza A result on the Hologic assay across this time period was 27.6 with a standard deviation of 5.9 (Figure S2). Influenza virus whole genome sequencing data was available for 109 of these specimens, and all contained seasonal influenza A virus subtype H3N2 (n=69) or subtype H1N1 (n=40) (Table S3).

At least one sample was available for 43 of 50 weeks across the time period studied for retrospective H5 subtyping (Figure 3). The median number of samples subtyped per week was 2 (IQR: 1-6). During the winter influenza season (epidemiological weeks 2024-51 to 2025-08), a median of 44 samples per week were subtyped with 512 samples in total. None of the samples tested positive for the H5 target (Table 6). The vast majority of samples (98.8%) tested positive for the pan-influenza A virus M target.

**Table 6:** H5 testing of influenza-A positive samples. Retrospective Subtyping refers to residual specimens tested for surveillance purposes. Routine Clinical Care refers to H5 subtyping tests ordered prospectively once the validated clinical assay was implemented.

| Specimen Type | N | H5 Positive | M Positive | Extraction Control Positive |
| --- | --- | --- | --- | --- |
| Retrospective Subtyping | 590 | 0 | 583 | 590 |
| Routine Clinical Care | 85 | 0 | 70 | 85 |

The patient population from whom these samples were collected (Table 7) consisted predominantly of adults, with a median age of 40 years (IQR: 26–59). Most samples were collected in emergency departments (n=379) or outpatient clinics (n=150). 18 samples were sourced from outside of the of the UW Medicine health system. Based on the address provided at the time of clinical testing, approximately 92% of all samples were obtained from patients residing in Seattle and Western Washington (Table S4) based on the first three digits of patients’ home zip code. Of 590 patients, 388 came from Seattle’s urban core. An additional 156 patients came from the greater Seattle metropolitan area or Western Washington. Beyond Western Washington, 12 samples came from patients residing in other parts of Washington State or Alaska. The remaining 26 samples originated from patients with homes addresses across the United States.

**Table 7:** Demographic information related to samples tested retrospectively for H5 influenza virus. Demographic information was obtained from the electronic health record. “Not Available” includes data values “N/A”, “Declined to answer”, and “Unknown.”

|  | Retrospective Subtyping |  | Routine Clinical Care |  |
| --- | --- | --- | --- | --- |
|  | N | % | N | % |
| **All Samples** | **590** |  | **85** |  |
| **Age** |  |  |  |  |
| < 1 year | 4 | 0.7 | 0 | 0.0 |
| < 18 years | 66 | 11.2 | 3 | 3.5 |
| >= 18 years | 524 | 88.8 | 82 | 96.5 |
| Not Available | 0 | 0.0 | 0 | 0.0 |
| **Race** |  |  |  |  |
| American Indian or Alaska Native | 13 | 2.2 | 1 | 1.2 |
| Asian | 57 | 9.7 | 0 | 0.0 |
| Black or African-American | 94 | 15.9 | 5 | 5.9 |
| Native Hawaiian or Pacific Islander | 7 | 1.2 | 1 | 1.2 |
| White | 334 | 56.6 | 17 | 20.0 |
| Another Race | 23 | 3.9 | 0 | 0.0 |
| Not Available | 62 | 10.5 | 61 | 71.8 |
| **Ethnicity** |  |  |  |  |
| Hispanic or Latino | 76 | 12.9 | 1 | 1.2 |
| Not Hispanic or Latino | 455 | 77.1 | 23 | 27.1 |
| Not Available | 59 | 10.0 | 61 | 71.8 |
| **Sex** |  |  |  |  |
| Female | 291 | 49.3 | 35 | 41.2 |
| Male | 297 | 50.3 | 50 | 58.8 |
| Not Available | 2 | 0.3 | 0 | 0.0 |
| **Source** |  |  |  |  |
| Emergency Department | 379 | 64.2 | 56 | 65.9 |
| Inpatient | 43 | 7.3 | 29 | 34.1 |
| Outpatient | 150 | 25.4 | 0 | 0.0 |
| Outside Source (not from UW Medicine) | 18 | 3.1 | 0 | 0.0 |

### Prospective clinical subtyping of influenza A-positive samples in February 2025

The H5 assay was implemented at UW Medicine on February 6, 2025. 85 H5 subtyping tests were ordered on Influenza A virus-positive patients during routine clinical care from February 6, 2025 to February 28, 2025. None of these samples tested positive for H5 influenza virus. The majority (82.4%) tested positive for the pan-influenza A virus target M.

Across this 4-week period, the median number of H5 subtyping test orders was 4 per day (IQR: 2-5). The majority of test orders (n=56) came from emergency departments and the remainder were ordered for patients admitted to an inpatient service (n=29). This volume represents approximately 19% of the 300 influenza A-positive test results from emergency departments and 60% of the 47 influenza A-positive test results from inpatient services during this time period.

The median age of patients who received a H5 subtyping test as part of clinical care was 62 years old (IQR: 45-75), which is significantly older than patients in the retrospective subtyping cohort (p<0.01 by Wilcoxon rank sum test) (Table 7). There were also more samples collected from male patients in this cohort than in the retrospective subtyping cohort (58.8% vs 50.3%), although this difference was not statistically significant (p=0.19, Chi-squared test).

## Discussion

Here, we report the design, validation, and clinical implementation of an RT-qPCR assay to subtype H5 influenza A virus in nasopharyngeal, nasal, and conjunctival specimens. The assay is sensitive, with a limit of detection of 230 copies/mL, and specific, with no cross-reactivity to common respiratory pathogens, including other influenza A virus subtypes. Recently, several other groups have validated and implemented laboratory developed tests for H5 subtyping, though little published performance characteristic data are available for comparison (18–20). The sensitivity of our assay is comparable to that of the assay recently published by Sahoo et al., with both assays reporting limits of detection below 5 copies per reaction (18). It is difficult to compare these values to the limit of detection reported for the CDC’s H5 subtyping kit, ∼250 EID50/mL, as this value is reported in units of egg infectious dose rather than absolute copy number. In both tissue culture models (31) and human infections (32), the number of template molecules detectable by RT-qPCR does not consistently correlate with viral titer because the ratio of non-infectious viral particles varies greatly between individual infections. However, when comparing the same templates side-by-side, the two assays displayed similar performance.

Given the preponderance of conjunctivitis reported in human cases related to the current outbreak, we validated the assay’s compatibility with conjunctival swab specimens. Although the CDC H5 subtyping kit is authorized for use with conjunctival swab specimens (33), to the best of our knowledge, no published validation studies have included this specimen type. We also validated the assay’s ability to detect both circulating genotypes that have caused human cases, B3.13 and D1.1.

Following validation, we tested 675 specimens from UW Medicine collected between March 2024 and February 2025 and identified no infections with subtype H5 virus. Importantly, we used the test to distinguish between H5 influenza virus and seasonal influenza viruses during the 2024-2025 influenza virus season, and influenza virus sequencing data was available for 109 of these specimens. This is the first surveillance study performed during a period of high seasonal influenza virus activity (30) since the outbreak began, and complements a recent study by Adams et al. that detected no H5N1 infections during a period of low seasonal influenza virus activity (34). Our dataset includes both retrospective testing of residual specimens (n=590) and clinical testing ordered during routine medical care after the assay was implemented at UW Medicine (n=85). This surveillance was performed in Washington State, which to date has experienced the second-highest number of human cases of H5N1 influenza infection.

Our results come with several limitations. First, the assay is designed to maximize sensitivity of the H5 targets at the expense of the M target. Because of this, the M target is less sensitive, and we have observed false-negative results on samples that are known to be influenza A-positive. This limitation is ameliorated by the ability of all influenza tests to detect H5N1 as influenza virus, and the indication of the subtyping assay to be used with known influenza-positive specimens. Second, the probe “H5 Probe 1” carries mismatches to the majority of recent H5 sequences from North America. This occurred because initial material for validation was extremely limited, and probes were designed to bind the only inactivated H5N1 reference material available (BEI NR-59421), which encoded an *HA* sequence from 2009. Since that time, updated inactivated H5N1 reference material (BEI NR-59886) has become available and was used for validation, highlighting the importance of making inactivated viral material available that can be used for BSL-2 validation activities. While our probe functions with contemporary H5N1 genotypes, future evolution might diminish probe binding and necessitate a revised probe sequence. Finally, although our surveillance data represent the largest set of influenza A-positive specimens tested for H5 in the literature since the outbreak began, almost all tested specimens tested originated from Seattle, Washington. Sampling from more rural regions, where patients are more likely to have exposure to livestock or avian species, is critical for both rapid detection of zoonotic infections and assessing point prevalence in a potentially higher risk region. With engagement and collaboration with clinicians and public health departments in rural areas, the clinically reportable H5 subtyping assay described here can help in these efforts.

Our data are most useful for understanding the prevalence of H5 infections during the 2024-2025 annual influenza virus season in a single academic medical system. We detected no cases of subtype H5 infection within the population of patients infected with influenza A viruses. As such, we did not detect evidence of any evolutionary events that can occur in the presence of co-circulation, such as recombination between H5 viruses and seasonal influenza A viruses (1). Our conclusions are in agreement with data from both the national influenza surveillance program, which has identified very few cases through non-specific respiratory disease surveillance (7), and the recent Adams et al. study, which found no H5 cases by screening clinical respiratory samples collected outside of the annual influenza season (34). Taken together, these data support current public health guidance indicating that H5 infection risk remains almost entirely associated with exposure to infected animals. Focused testing of patients with epidemiological risk factors, such as farm workers, and prompt treatment with antiviral medications remain cornerstones of clinical management (35). However, as H5N1 influenza virus is expected to spread in animal species in the United States for the foreseeable future, integration of routine H5 testing into health systems remains critical to monitor for future changes to the epidemiology of this outbreak.

## Data availability

Viral sequences referenced in this manuscript are available under BioProject PRJNA1232193.

## References

1. Kandeil A, Patton C, Jones JC, Jeevan T, Harrington WN, Trifkovic S, et al. Rapid evolution of A(H5N1) influenza viruses after intercontinental spread to North America. Nat Commun [Internet]. 2023 May 29 [cited 2025 Mar 4];14(1):3082. Available from: https://www.nature.com/articles/s41467-023-38415-7

2. Preliminary report on genomic epidemiology of the 2024 H5N1 influenza A virus outbreak in U.S. cattle (Part 1 of 2) - Influenza virus / H5N1-global [Internet]. Virological. 2024 [cited 2024 Aug 5]. Available from: https://virological.org/t/preliminary-report-on-genomic-epidemiology-of-the-2024-h5n1-influenza-a-virus-outbreak-in-u-s-cattle-part-1-of-2/970

3. Peacock TP, Moncla L, Dudas G, VanInsberghe D, Sukhova K, Lloyd-Smith JO, et al. The global H5N1 influenza panzootic in mammals. Nature [Internet]. 2025 Jan [cited 2025 Feb 1];637(8045):304–13. Available from: https://www.nature.com/articles/s41586-024-08054-z

4. Ison MG, Marrazzo J. The emerging threat of H5N1 to human health. N Engl J Med [Internet]. 2024 Dec 31 [cited 2025 Jan 3]; Available from: https://www.nejm.org/doi/full/10.1056/NEJMe2416323

5. APHIS Confirms D1.1 Genotype in Dairy Cattle in Nevada [Internet]. Animal and Plant Health Inspection Service. [cited 2025 Feb 10]. Available from: https://www.aphis.usda.gov/news/program-update/aphis-confirms-d11-genotype-dairy-cattle-nevada-0

6. Highly Pathogenic Avian Influenza A (H5N1) Virus Infection Reported in a Person in the U.S [Internet]. CDC. 2024 [cited 2024 Jul 29]. Available from: https://www.cdc.gov/media/releases/2024/p0401-avian-flu.html

7. CDC. H5 Bird Flu: Current Situation [Internet]. Avian Influenza (Bird Flu). 2025 [cited 2025 Feb 10]. Available from: https://www.cdc.gov/bird-flu/situation-summary/index.html

8. Jassem AN, Roberts A, Tyson J, Zlosnik JEA, Russell SL, Caleta JM, et al. Critical illness in an adolescent with influenza A(H5N1) virus infection. N Engl J Med [Internet]. 2024 Dec 31 [cited 2025 Feb 1]; Available from: https://www.nejm.org/doi/full/10.1056/NEJMc2415890

9. LDH reports first U.S. H5N1-related human death [Internet]. [cited 2025 Feb 1]. Available from: https://ldh.la.gov/news/H5N1-death

10. Garg S, Reinhart K, Couture A, Kniss K, Davis CT, Kirby MK, et al. Highly pathogenic avian influenza A(H5N1) virus infections in humans. N Engl J Med [Internet]. 2024 Dec 31 [cited 2025 Feb 1]; Available from: https://www.nejm.org/doi/full/10.1056/NEJMoa2414610

11. Mellis AM, Coyle J, Marshall KE, Frutos AM, Singleton J, Drehoff C, et al. Serologic evidence of recent infection with highly pathogenic avian influenza A(H5) virus among dairy workers - Michigan and Colorado, June-August 2024. MMWR Morb Mortal Wkly Rep [Internet]. 2024 Nov 7 [cited 2025 Mar 1];73(44):1004–9. Available from: https://www.cdc.gov/mmwr/volumes/73/wr/mm7344a3.htm

12. Health Alert Network (HAN) - 00520 [Internet]. 2025 [cited 2025 Feb 10]. Available from: https://www.cdc.gov/han/2025/han00520.html

13. CDC. Interim Guidance on the Use of Antiviral Medications for Treatment of Human Infections with Novel Influenza A Viruses Associated with Severe Human Disease [Internet]. Avian Influenza (Bird Flu). 2024 [cited 2025 Feb 10]. Available from: https://www.cdc.gov/bird-flu/hcp/novel-av-treatment-guidance/index.html

14. CDC. Interim Guidance for Infection Control Within Healthcare Settings When Caring for Confirmed Cases, Probable Cases, and Cases Under Investigation for Infection with Novel Influenza A Viruses Associated with Severe Disease [Internet]. Avian Influenza (Bird Flu). 2024 [cited 2025 Feb 10]. Available from: https://www.cdc.gov/bird-flu/hcp/novel-flu-infection-control/index.html

15. Pinsky BA, Bradley BT. Opportunities and challenges for the U.S. laboratory response to highly pathogenic avian influenza A(H5N1). J Clin Virol. 2024 Oct;174(105723):105723.

16. Bedford T, Greninger AL, Roychoudhury P, Starita LM, Famulare M, Huang M-L, et al. Cryptic transmission of SARS-CoV-2 in Washington state. Science [Internet]. 2020 Oct 30 [cited 2025 Feb 11];370(6516):571–5. Available from: https://pmc.ncbi.nlm.nih.gov/articles/PMC7810035/

17. 06/12/2024: Lab Advisory: CDC Open Call to Industry for Influenza A(H5) Diagnostic Test Development and Validation [Internet]. 2024 [cited 2024 Jul 29]. Available from: https://www.cdc.gov/locs/2024/06-12-2024-Lab-Advisory_CDC-Open-Call-Industry-Influenza-AH5-Diagnostic-Test-Development-Validation.html

18. Sahoo MK, Morante IEA, Huang C, Solis D, Yamamoto F, Ohiri UC, et al. Multiplex dual-target reverse transcription PCR for subtyping avian influenza A(H5) virus. Emerg Infect Dis [Internet]. 2024 Jul 10 [cited 2024 Jul 14];30(8). Available from: https://wwwnc.cdc.gov/eid/article/30/8/24-0785_article

19. Quest Diagnostics Offers Test to Detect Avian Influenza Virus [Internet]. Quest Diagnostics Newsroom. [cited 2025 Feb 28]. Available from: https://newsroom.questdiagnostics.com/press-releases?item=94382

20. Labcorp Launches H5 Bird Flu Test in the U.S., Now Available for Order through Physicians [Internet]. Labcorp. [cited 2025 Feb 28]. Available from: https://ir.labcorp.com/news-releases/news-release-details/labcorp-launches-h5-bird-flu-test-us-now-available-order-through

21. CDC. Update: Human Infection with Highly Pathogenic Avian Influenza A(H5N1) Virus in Texas [Internet]. National Center for Immunization and Respiratory Diseases. 2024 [cited 2025 Feb 5]. Available from: https://www.cdc.gov/ncird/whats-new/human-infection-H5N1-bird-flu.html

22. Kuypers J, Wright N, Ferrenberg J, Huang M-L, Cent A, Corey L, et al. Comparison of real-time PCR assays with fluorescent-antibody assays for diagnosis of respiratory virus infections in children. J Clin Microbiol [Internet]. 2006 Jul [cited 2025 Feb 24];44(7):2382–8. Available from: https://pubmed.ncbi.nlm.nih.gov/16825353/

23. Sayers EW, Bolton EE, Brister JR, Canese K, Chan J, Comeau DC, et al. Database resources of the national center for biotechnology information. Nucleic Acids Res [Internet]. 2022 Jan 7 [cited 2025 Feb 6];50(D1):D20–6. Available from: https://pubmed.ncbi.nlm.nih.gov/34850941/

24. Katoh K, Standley DM. MAFFT multiple sequence alignment software version 7: improvements in performance and usability. Mol Biol Evol [Internet]. 2013 Apr 16 [cited 2025 Jan 16];30(4):772–80. Available from: https://academic.oup.com/mbe/article-pdf/30/4/772/6420419/mst010.pdf

25. Metzger C, Lienhard R, Seth-Smith H, Roloff T, Wegner F, Sieber J, et al. PCR performance in the SARS-CoV-2 Omicron variant of concern? Schweiz Med Wochenschr [Internet]. 2021 Dec 6 [cited 2025 Feb 3];151(4950):w30120. Available from: https://smw.ch/index.php/smw/article/view/3124/5214

26. FDA. Class 2 Device Recall CDC, Influenza A/H5 Subtyping Kit [Internet]. [cited 2025 Feb 3]. Available from: https://www.accessdata.fda.gov/scripts/cdrh/cfdocs/cfres/res.cfm?id=208126

27. WHO. Information for molecular diagnosis of influenza virus, 7th revision [Internet]. 2021 [cited 2024 Aug 7]. Available from: https://www.who.int/teams/global-influenza-programme/laboratory-network/quality-assurance/eqa-project/information-for-molecular-diagnosis-of-influenza-virus

28. CDC. CDC’s Influenza SARS-CoV-2 Multiplex Assay [Internet]. Centers for Disease Control and Prevention. 2023 [cited 2025 Feb 3]. Available from: https://archive.cdc.gov/www_cdc_gov/coronavirus/2019-ncov/lab/multiplex.html

29. Bruce EA, Huang M-L, Perchetti GA, Tighe S, Laaguiby P, Hoffman JJ, et al. Direct RT-qPCR detection of SARS-CoV-2 RNA from patient nasopharyngeal swabs without an RNA extraction step. PLoS Biol [Internet]. 2020 Oct;18(10):e3000896. Available from: 10.1371/journal.pbio.3000896

30. Pacific Northwest Respiratory Virus Epidemiology Data [Internet]. [cited 2025 Mar 7]. Available from: https://depts.washington.edu/labmed/pnw_respiratory_dashboard/

31. Xue J, Chambers BS, Hensley SE, López CB. Propagation and characterization of influenza virus stocks that lack high levels of defective viral genomes and hemagglutinin mutations. Front Microbiol [Internet]. 2016 Mar 23 [cited 2024 Aug 7];7:326. Available from: https://www.frontiersin.org/journals/microbiology/articles/10.3389/fmicb.2016.00326/pdf

32. Vasilijevic J, Zamarreño N, Oliveros JC, Rodriguez-Frandsen A, Gómez G, Rodriguez G, et al. Reduced accumulation of defective viral genomes contributes to severe outcome in influenza virus infected patients. PLoS Pathog [Internet]. 2017 Oct 12 [cited 2024 Aug 7];13(10):e1006650. Available from: https://journals.plos.org/plospathogens/article/file?id=10.1371/journal.ppat.1006650&type=printable

33. CDC. 05/31/2024 Enforcement Discretion Granted for the Use of Conjunctival Swabs with the CDC Human Influenza Virus Real-Time RT-PCR Diagnostic Panel, Influenza A/H5 Subtyping Kit [Internet]. Laboratory Outreach Communication System (LOCS). 2024 [cited 2025 Mar 4]. Available from: https://www.cdc.gov/locs/php/messages/2024/2024-05-31-enforcement-discretion-influenza-ah5-subtyping-kit.html

34. Adams GC, Devlin JE, Klontz E, Laing RA, Branda JA, Chowdhury N, et al. Combing the haystacks: The search for highly pathogenic avian influenza virus using a combined clinical and research-developed testing strategy [Internet]. medRxiv. 2025 [cited 2025 Mar 4]. p. 2025.02.12.25321810. Available from: https://www.medrxiv.org/content/10.1101/2025.02.12.25321810v1.abstract

35. CDC. Highly Pathogenic Avian Influenza A(H5N1) Virus: Interim Recommendations for Prevention, Monitoring, and Public Health Investigations [Internet]. Avian Influenza (Bird Flu). 2025 [cited 2025 Mar 4]. Available from: https://www.cdc.gov/bird-flu/prevention/hpai-interim-recommendations.html

