## Supplementary material for "Validation of H5 influenza virus subtyping RT-qPCR assay and low prevalence of H5 detection in 2024-2025 influenza virus season": Figure S1

| **Oligo Name** | **H5 Forward 1** | **H5 Probe 1** | **H5 Reverse 1** |
|---|---|---|---|
| **Oligo Sequence** | TGGAAAGTGTGAGAAATGGGACGT | TGACTACCCGCAGTATTCAGAAGAAGCAAGACTAA | CAGCGGCAAGTTCCCTAGCA |
| **NIST HA** | ........................ | .........T.....................T... | .................... |
| **B3.13 HA** | ........................ | .........T.....................T... | .................... |
| **D1.1 HA** | ........................ | .........T....................GT... | .......G............ |

| **Oligo Name** | **H5 Forward 2** | **H5 Probe 2** | **H5 Reverse 2** |
|---|---|---|---|
| **Oligo Sequence** | TGGGTACCATCATAGCAATGAGCA | TGGGTACGCTGCGGACAAAGAATCCA | TTTGAGGCAGTTGGAAGGGAGTT |
| **NIST HA** | ........................ | .......................... | ....................... |
| **B3.13 HA** | ........................ | .......................... | ....................... |
| **D1.1 HA** | ........................ | ............A............. | ....................... |

**Figure S1: H5 Primer and probe binding sites.**
NIST RNA refers to the sequence of the *HA* gene in the NIST RNA template (GenBank: N90JTPPO). B3.13 HA (GenBank: PP692142) and D1.1 HA (GenBank: PQ585632) are representative sequences of each genotype.
