## Supplementary material for "Validation of H5 influenza virus subtyping RT-qPCR assay and low prevalence of H5 detection in 2024-2025 influenza virus season": Figure S2

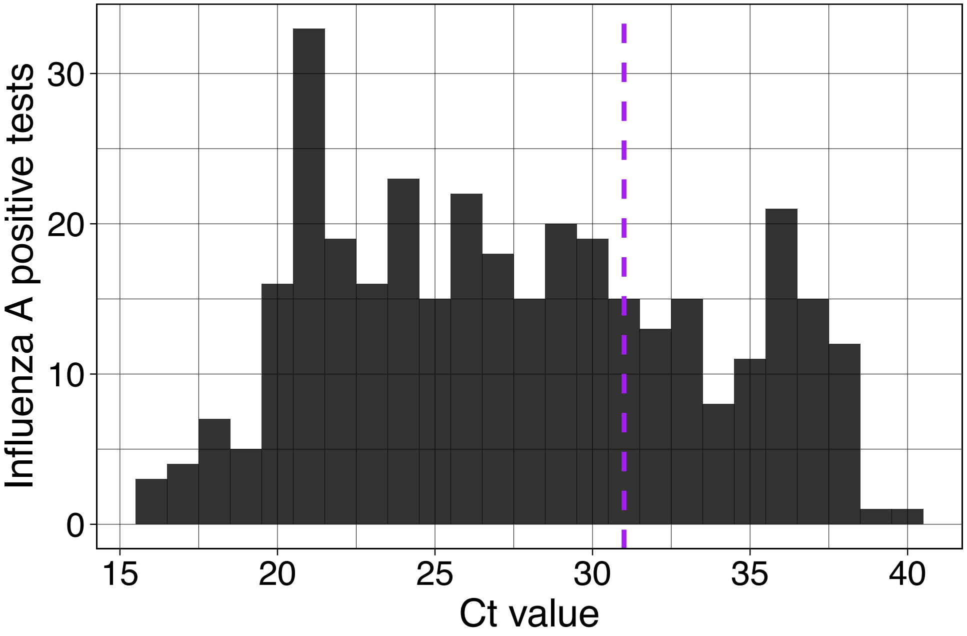


**Figure S2: Ct values of influenza-positive Hologic Panther Fusion Flu A/B/RSV assay results collected between March 2024 and February 2025.** (n=348) Dashed line set at Ct value 31, which is the threshold for sample collection for genomic surveillance. Residual genomic surveillance samples were used for retrospective subtyping in this study.
